# Bone tracers for transthyretin amyloid cardiomyopathy: are [^99m^Tc]Tc-DPD and [^99m^Tc]Tc-HMDP truly equivalent?

**DOI:** 10.1101/2024.02.14.24302851

**Authors:** Julien Dubois, Florentin Kucharczak, Denis Mariano-Goulart, Tom Paunet

## Abstract

**Background:** The management of transthyretin amyloid cardiomyopathy (ATTR-CM) has revolved around the scintigraphic diagnosis since the introduction of a specific treatment; however, the equivalency of the bone radiotracers remains unclear. This retrospective monocentric observational study compared [^99m^Tc]Tc-3,3-diphosphono-1,2-propanodicarboxylic acid ([^99m^Tc]Tc-DPD) and [^99m^Tc]Tc-hydroxy-methylene diphosphonate ([^99m^Tc]Tc-HMDP) for ATTR-CM diagnosis.

**Methods:** One hundred and twenty-nine patients who underwent single photon emission computed tomography (SPECT/CT) after intravenous injection of [^99m^Tc]Tc-DPD or [^99m^Tc]Tc-HMDP for ATTR-CM were included. The patients’ current visual Perugini grades were retrieved. Regions of interest (ROI) were measured on the heart (H) and on contralateral mediastinum (CM), and H/CM ratios were calculated.

**Results:** Although the distribution of quantitative assessments of heart to contralateral mediastinum ratios is wider with [^99m^Tc]Tc-DPD, suggesting a trend towards improved diagnosis, no difference in Perugini grades was found between [^99m^Tc]Tc-DPD or [^99m^Tc]Tc-HMDP for the diagnosis of ATTR-CM in evocative/non-evocative conditions. There was no difference in ATTR-CM diagnosis between the 2 tracers with a threshold of 1.5 (p-value = 3.316*10^−10^ for [^99m^Tc]Tc-HMDP and p-value = 2.59*10^−15^ for [^99m^Tc]Tc-DPD).

**Conclusions:** We show in our local cohort that [^99m^Tc]Tc-DPD and [^99m^Tc]Tc-HMDP for amyloidosis diagnostic are equivalent for ATTR-CM diagnosis based on the Perugini grading scale. With [^99m^Tc]Tc-DPD, a wider range of the H/CM ratio was noted, which may be considered as a better discrimination of the disease by this bone tracer. Additional research with a final diagnosis of the disease is necessary to evaluate the utility of this quantitative evaluation.

## Introduction

Bone tracers’ scintigraphy have long been used for transthyretin amyloid cardiomyopathy (ATTR-CM) diagnosis complement. If the diagnosis is suspected based on typical symptoms or abnormal cardiac test results, the gold standard to confirm the diagnosis is endomyocardial biopsy. This may be supplemented by ^99m^Tc-bisphosphonate single-photon emission computed tomography (SPECT), cardiac magnetic resonance imaging (MRI) or possibly a genetic test to differentiate wild type amyloidosis and hereditary amyloidosis. Noninvasive techniques allow the diagnosis of ATTR-CM to be confirmed without the need for an invasive procedure and prevail in worldwide guidelines [1–4]. ATTR-CM remains an under-diagnosed disease, but progress continues to be made in understanding its pathological process, enabling early diagnosis and the development of treatments [5]. Thus, the mechanism behind bone tracer uptake in ATTR-CM is still unclear. Hypothesis suggest the presence of microcalcifications that are more common in ATTR-CM than in amyloid light chain cardiomyopathy (AL-CM), which could led to a higher bone tracer uptake [6–8]. There must be sufficient accumulation of amyloid in the myocardium for ^99m^Tc-bisphosphonate scintigraphy to be positive, and this seems to depend on the composition of the amyloid fibrils in transthyretin [9,10].

[^99m^Tc]Tc-3,3-diphosphono-1,2-propanodicarboxylic acid ([^99m^Tc]Tc-DPD), [^99m^Tc]Tc-hydroxy-methylene diphosphonate ([^99m^Tc]Tc-HMDP) and [^99m^Tc]Tc-pyrophosphate ([^99m^Tc]Tc-PYP) are the three SPECT bone tracers approved for assessing ATTR-CM diagnostic [11,12]. [^99m^Tc]Tc-PYP is not recommended in Europe for amyloidosis exploration, although it is preferentially used in the United States of America [13]. Not all ^99m^Tc-bisphosphonates necessarily have the same sensitivity for detecting ATTR-CM, and some authors do not recommend the use of [99mTc]Tc-methylene diphosphonate ([^99m^Tc]Tc-MDP) in this indication [6]. However, the latest reviews and meta-analysis suggest that there is no real difference in ATTR-CM diagnostic performance between the 3 indicated tracers [14–17]. Abulizi and al. show comparable myocardial uptake intensity of [^99m^Tc]Tc-HMDP and [^99m^Tc]Tc-DPD on early-phase scintigraphy in matched patients with hereditary ATTR-CM [15].

Recently, our center switched from using [^99m^Tc]Tc-DPD exclusively for amyloidosis to [^99m^Tc]Tc-HMDP for financial and literary justifications. Our retrospective study uses the populations in our database before and after this change to evaluate the hypothesis of equivalence of these two radiotracers in the diagnosis of ATTR-CM based on the Perugini classification [6,12,18].

## Methods

### Patients

Single-center retrospective observational study has been conducted in the nuclear medicine department of the Montpellier University Hospital for two years, from December 2021 to November 2023. All patients underwent late-phase bone scan after intravenous injection of [^99m^Tc]Tc-DPD or [^99m^Tc]Tc-HMDP for an ATTR-CM suspicion. Patients were recruited and separated in two arms depending on the radiotracer used. Parameters gathered were gender, age, weight, and injected dose. Patients’ characteristics are presented in Table 1.

**Table 1:** Patient characteristics *MBq: MegaBecquerel; H/CM: heart/contralateral mediastinum; SD: Standard deviation*.

| Characteristics | $^{99m}\text{Tc}$[Tc-HMDP,<br>N = 61<sup>1</sup> | $^{99m}\text{Tc}$[Tc-DPD,<br>N = 68<sup>1</sup> | p-value |
| --- | --- | --- | --- |
| **Gender** |  |  | 0.9<sup>2</sup> |
| *F* | 26 (43%) | 30 (44%) |  |
| *M* | 35 (57%) | 38 (56%) |  |
| **Age (years)** | 78 (10) | 77 (12) | 0.9<sup>3</sup> |
| **Activity (MBq)** | 706 (127) | 712 (115) | 0.8<sup>3</sup> |
| **Perugini score** |  |  | 0.2<sup>4</sup> |
| *0* | 48 (79%) | 47 (69%) |  |
| *1* | 4 (6%) | 5 (7.5%) |  |
| *2* | 9 (15%) | 11 (16%) |  |
| *3* | 0 (0%) | 5 (7.5%) |  |
| **H/CM ratio** | 1.20 (0.32) | 1.35 (0.66) | 0.8<sup>3</sup> |

<sup>1</sup> n (%); Mean (SD)
<sup>2</sup> Pearson's Chi-squared test
<sup>3</sup> Wilcoxon rank sum test
<sup>4</sup> Fisher's exact test

This study was conducted in accordance with the Helsinki Declaration and was approved by the French nuclear medicine committee for the protection of individuals (CEMEN) the 05/12/2023 and registered under the institutional review board number CEMEN-2023-04. Written informed consent was obtained by all the patients for publication and accompanying images.

### Bone scan

All patients underwent late-phase bone scan 3 hours after [^99m^Tc]Tc-DPD or [^99m^Tc]Tc-HMDP injection. Whole-body scan in the anterior and posterior views were acquired using a dual-headed SPECT/CT camera (Discovery NM 870, GE HealthCare, Chicago, USA) using low energy, high resolution collimators, at 20 cm/min speed, with 256×256 matrix [12].

### Usual scintigraphy interpretation

In the investigation of cardiac amyloidosis (especially ATTR-CM), the Perugini grading scale is a semi-quantitative technique for assessing cardiac uptake after bone tracer scintigraphy. Tracer uptake in the heart and ribs is visually compared using a grading system ranging from 0 to 3. Interpretations that receive a visual grade of two or three on scintigraphy imaging are categorized as ATTR positive. Perugini grading score 0 and 1 are considered ATTR negative [6,12]. The Perugini grades of each patient were obtained from subjective interpretations made by senior nuclear physicians.

### Proposed metric for assessing continuous uptake levels

We intend to determine if distribution of the heart to contralateral mediastinum ratios varies depending on the two tracers used. The measurements are based on the geometric means of whole-body images and represented in Figure 1. If pathological activity was visible in the cardiac area, a Region of Interest (ROI) was delineated around it. The background ROI in the contralateral mediastinum (CM) was created by horizontally symmetrizing the heart (H) ROI with respect to the median axis of the sternal manubrium (Figure 1a). If no pathological activity was visible in the cardiac area, a circular H ROI with a diameter of 60% of the width of the hemi-thorax was drawn in the cardiac region. The CM ROI was created using the same procedure as mentioned earlier, employing horizontal symmetry of the cardiac ROI with respect to the axis of the sternal manubrium (Figure 1b). In both cases, if an intense focal uptake, unrelated to amyloidosis, was present within any of the ROIs, it was excluded (e.g., rib fracture, bone metastasis). The heart ratio of each subject (H/CM) was calculated based on the average activities within each ROI [19].

**Figure 1:**
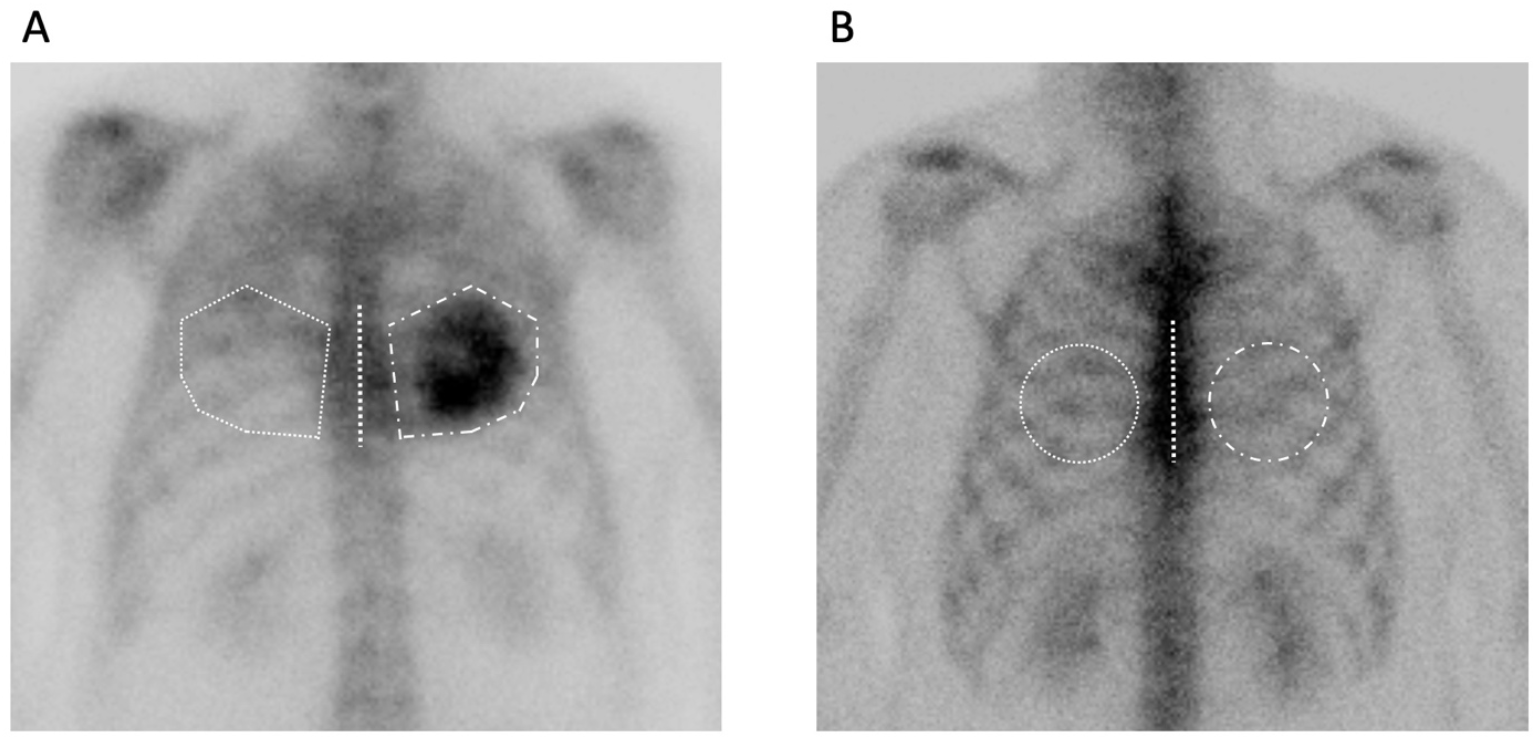
Representative heart/contralateral mediastinum (H/CM) ratio measurements. *(A) If pathological activity was visible in the cardiac area, the background region of interest (ROI) in the contralateral mediastinum was created by horizontally symmetrizing the heart ROI with respect to the median axis of the sternal manubrium; (B) If no pathological activity was visible in the cardiac area, a circular heart ROI with a diameter of 60% of the width of the hemi-thorax was drawn in the cardiac region, and the contralateral mediastinum ROI was created by horizontally symmetrizing the heart ROI with respect to the median axis of the sternal manubrium*.

### Data management and statistical analysis

All data were recovered as described by Good Clinical Practices (GCP) guidelines [20]. Epidemiological, clinical, paraclinical, biological, pharmaceutical, and radiopharmaceutical dispensing data were collected in patients record and anonymized. H/CM ratios were computed using the open-source software ITK-SNAP version 4.0 (Penn Image Computing and Science Laboratory) [21]. The Wilcoxon test was performed to compare our groups. Statistical analyses were performed using R^®^ software 4.1.3. Statistical significance was defined as *P* ≤ 0.05.

## Results

One hundred and twenty-nine patients with ATTR-CM suspicion were recruited: 61 in the [^99m^Tc]Tc-HMDP group and 68 in the [^99m^Tc]Tc-DPD group. There was no significant difference between the two groups in terms of age and gender. The mean injected dose was 9.9 +/-0.86 MBq.kg^-1^ (0.27 +/-0.023 mCi.kg^-1^).

No statistical differences in Perugini grades and in mean H/CM ratio were found between our two groups as depicted in table 1.

H/CM ratio’s distribution and probability density gave a first descriptive aspect of the bisphosphonate performance with more 1:1 ratio obtained for [^99m^Tc]Tc-HMDP, and more 1:2 and higher ratio for [^99m^Tc]Tc-DPD (Figure 2). [^99m^Tc]Tc-DPD group had more ratios below 1 (n=18) than [^99m^Tc]Tc-HMDP group (n = 11). Ten patients who underwent the exploration with [^99m^Tc]Tc-DPD had a H/CM ratio superior to the maximum H/CM ratio obtained with [^99m^Tc]Tc-HMDP.

**Figure 2:**
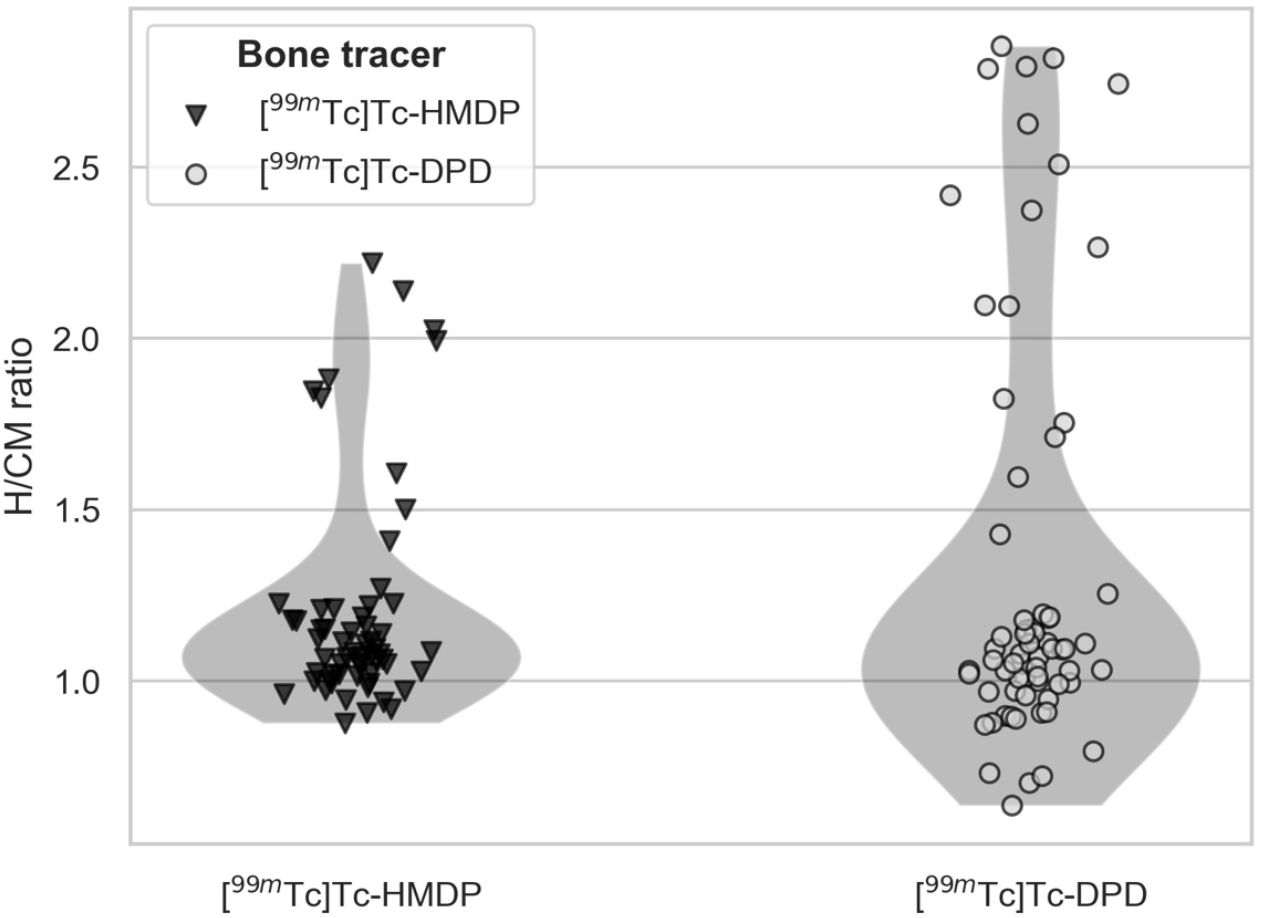
Heart/contralateral mediastinum (H/CM) ratio’s distribution between [^99m^Tc]Tc-HMDP (n=61) and [^99m^Tc]Tc-DPD (n=68)

By combining these results with the Perugini grading scores found for each patient, we obtained a heterogeneous distribution of related H/CM ratios (Figure 3). We pointed out that grade 0 and grade 1 Perugini scored patients are comparable in their mean H/CM ratio, with a H/CM ratio threshold of 1.5 for [^99m^Tc]Tc-DPD and [^99m^Tc]Tc-HMDP (p-value = 0.17, and p-value = 0.08 respectively). One patient had a H/CM ratio of 1.502 associated to a Perugini score of 0. Ratios higher than 1.5 were related to Perugini grading score 2 and 3. Once we pooled 0 and 1 grading scores, and 2 and 3 grading scores, a statistically significant difference was observed for both bone tracers with p<0.001 (p-value = 1.431*10^−14^ for [^99m^Tc]Tc-DPD, and p-value = 2.253*10^−12^ for [^99m^Tc]Tc-HMDP). There was no difference in ATTR-CM diagnosis (considering the pooled Perugini groups as reference) between the 2 tracers with a threshold of 1.5 (p-value = 3.316*10^−10^, sensibility (Se) = 89% and specificity (Sp) = 98% for [^99m^Tc]Tc-HMDP; and p-value = 2.59*10^−15^, Se = 100% and Sp = 100% for [^99m^Tc]Tc-DPD). One patient was assessed Perugini 2 with a H/CM ratio below 1.5, due to low overall uptake. Another patient was assessed Perugini 3 despite having a H/CM ratio below 2, because he had diffuse amyloidosis with extraosseous tissue fixation. Representative images showing the less and the most heart uptake of bone tracers are shown in Figure 4.

**Figure 3:**
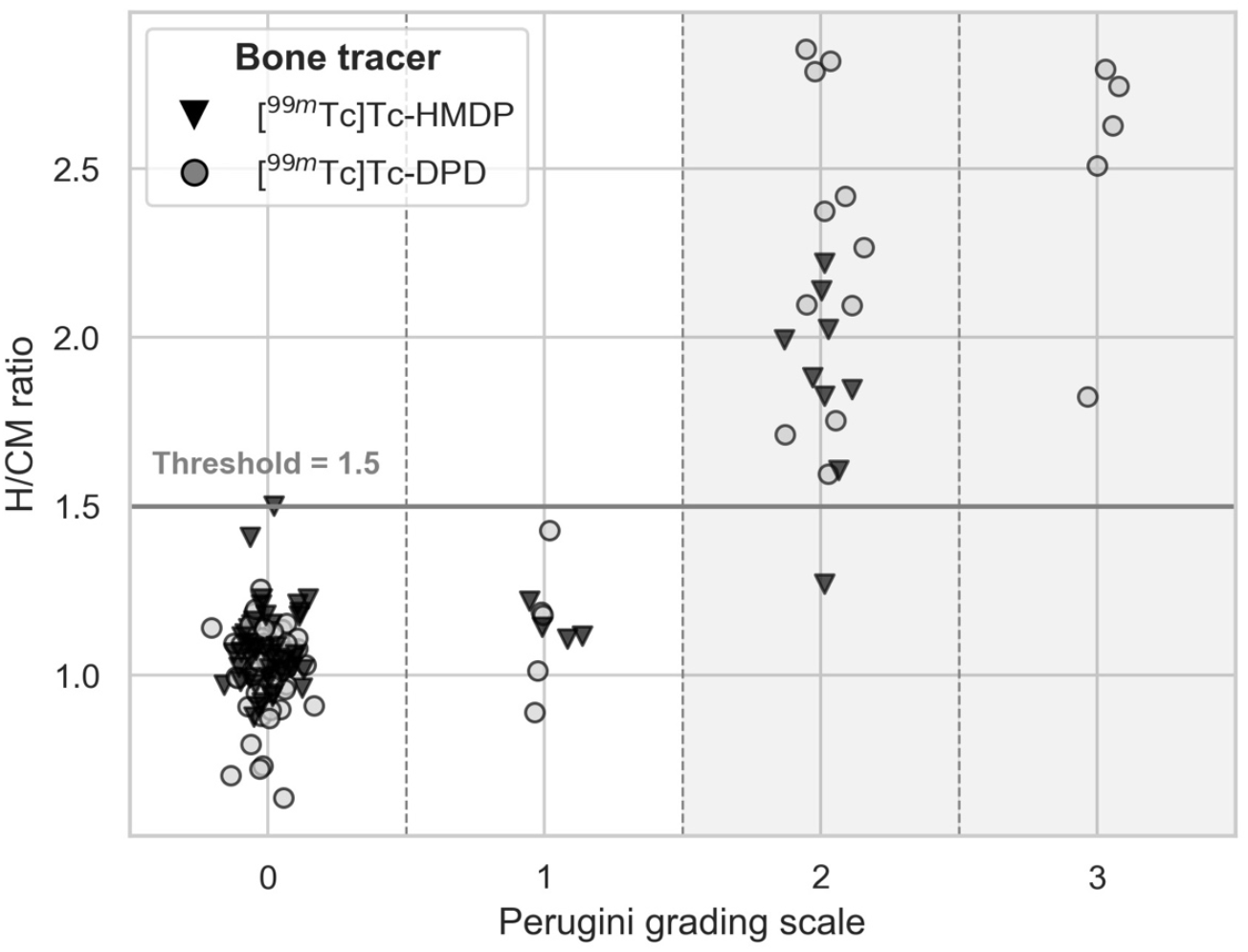
Heart/contralateral mediastinum (H/CM) ratio’s distribution according to attributed Perugini grading score for each patient

**Figure 4:**
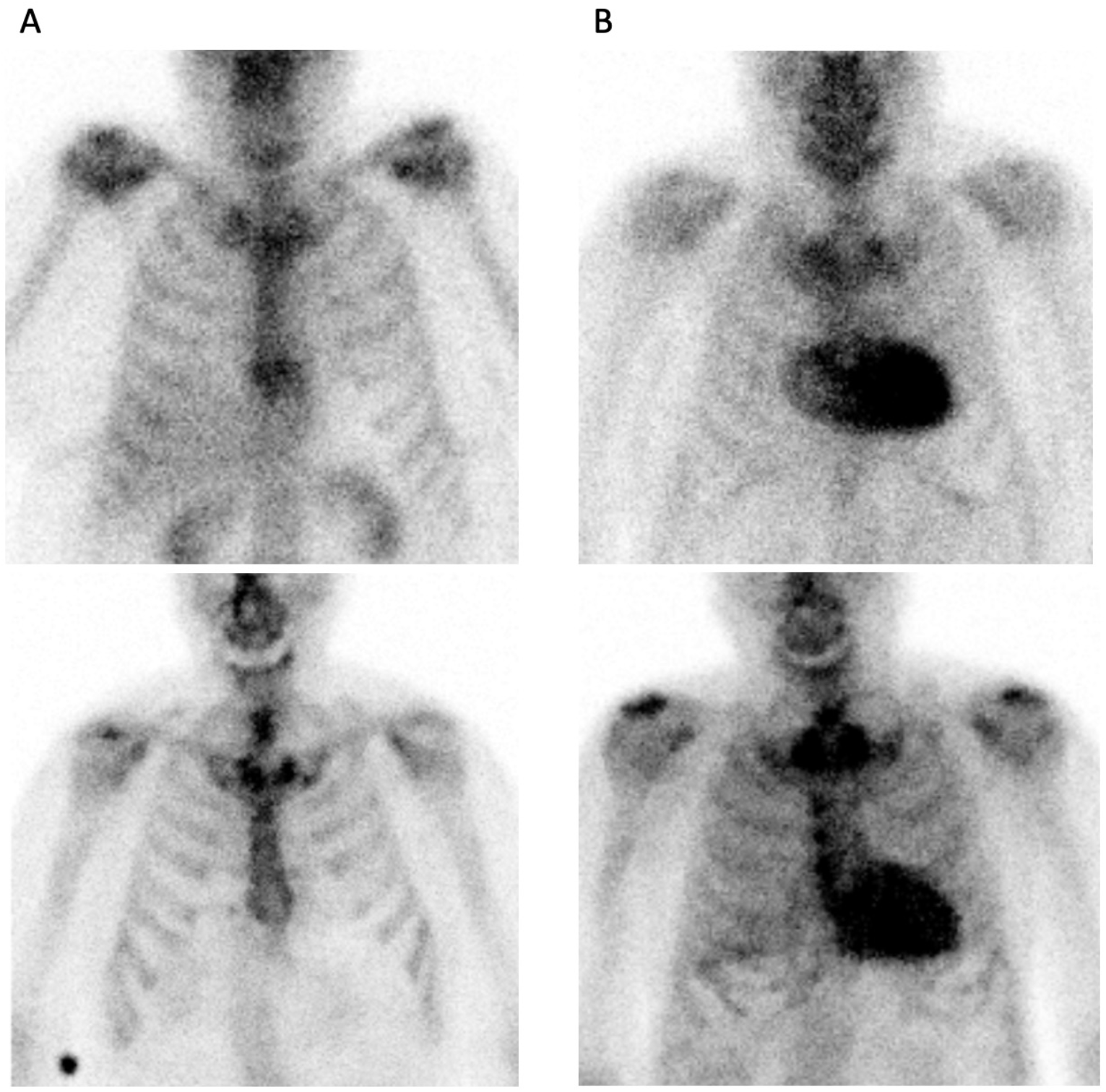
Representative images of (A) minimum uptake, and (B) maximum uptake of [^99m^Tc]Tc-DPD (up) and [^99m^Tc]Tc-HMDP (down). *Minimum H/CM ratio for [*^*99m*^*Tc]Tc-DPD and [*^*99m*^*Tc]Tc-HMDP were 0*.*64 and 0*.*88 respectively. Maximum H/CM ratio for [*^*99m*^*Tc]Tc-DPD and [*^*99m*^*Tc]Tc-HMDP were 2*.*85 and 2*.*22 respectively*.

## Discussion

In our cohort study of one hundred and twenty-nine patients, we showed that our two bone tracers are equivalent in terms of sensibility and specificity in evocative/non-evocative conditions for the diagnosis of ATTR-CM. We found no statistical difference between our two bone tracers in terms of H/CM ratios distribution, or Perugini grading scores distribution. However, when comparing [^99m^Tc]Tc-DPD to [^99m^Tc]Tc-HMDP, [^99m^Tc]Tc-DPD showed a larger range of H/CM ratios, from the 1.5 threshold both positive and negative, without consequences on the diagnostic of ATTR-CM. We have observed that the minimum and the maximum H/CM ratio are different for the two bone tracers in our cohort, which supports our last hypothesis.

In both hereditary and wild-type ATTR-CM, serial [^99m^Tc]Tc-DPD SPECT/CT imaging may be a useful tool for measuring and tracking response to disease-specific treatments [22]. Nevertheless, some hereditary ATTR-CM, like Phe64Leu mutation, appears to result in a lack of myocardial uptake of bone tracers [23]. A study showed that [^99m^Tc]Tc-HMDP uptake is significantly correlated with histological amyloid load in endomyocardial biopsy [24]. Also, an *in-vitro* study showed a higher affinity of [^99m^Tc]Tc-DPD for amyloid fibrils than [^99m^Tc]Tc-HMDP or [^99m^Tc]Tc-PYP [25]. Wollenweber and al. suggest that [^99m^Tc]Tc-DPD scintigraphy is useful to detect patients with a higher risk for polyneuropathy associated to ATTR-CM [26]. Regrettably, we were unable to assess the binding ratio of the two tracers concerning the causal mutation, as such investigations are exceedingly rare in the context of positive scintigraphy.

The specificity of ^99m^Tc-bisphophonate SPECT still need to be explored as studies show that chloroquine and derivatives mediated cardiotoxicity seem to induce false positive bone tracer uptake [27,28]. Few studies assessed the possibilities for using bone scintigraphy to monitor tafamidis therapy and found positive results for [^99m^Tc]Tc-DPD [29–32], but not conclusive for [^99m^Tc]Tc-PYP [33]. Mechanisms remain unclear and a systematic disease follow-up by bone tracer scintigraphy is therefore not recommended. There is also no specific positron emission tomography (PET) tracer to date, but several could be used for amyloidosis without distinguishing clearly cardiac amyloidosis subtypes [34]. Their uptake mechanisms also differ with SPECT bone tracer and their performance in the diagnosis of ATTR-CM remains to be demonstrated [35].

There is a degree of subjectivity in the visual interpretation of the Perugini grading score, particularly between scores 0 and 1, which are negative, and scores 2 and 3, which are positive. In our view, it is not possible to monitor disease or treatment simply using a Perugini grading score, but uptake quantification could be of interest in this context if the same tracer is administered and using the same acquisition modalities. Quantitative assessment could be an answer to this subjectivity by avoiding competition for the radiotracer with amyloid in bones, but still need to be prospectively studied [19,36,37]. As no statistical difference was found in meta-analyses, heterogeneity in methodology and patients across the studies may be a potential cause of bias impacting the quality of their results [38]. Prospective studies with higher quality inclusion criteria must be led to support one to another bone tracer, especially if hereditary mutation induce different uptake.

This single center, retrospective study focuses on a highly uncommon pathology that has a wide spectrum of physiopathology and treatment approach. All patient final diagnostics were not found in their medical record. We also have not enough information about treatment start; thus, we were not able to include this parameter in our analysis. The current work needs more confirmation because it is only the initial step toward future multicentric and prospective works. A larger cohort size with lower inter-patient variability will enable better management of data interpretation due to prospective patient inclusion. Homogeneity in group sizes, accurate patient’s amyloidosis classification, and better data collection management are also useful. Additionally, there may have been a temporal bias because our two groups followed each other chronologically after the radiotracer was switched.

In conclusion, our study shows that a diagnosis of ATTR-CM based on assessment of the Perugini score is not altered using [^99m^Tc]Tc-DPD or [^99m^Tc]Tc-HMDP. A wider range of the H/CM ratio was observed with [^99m^Tc]Tc-DPD and further studies with a final diagnosis of the disease are required to assess the value of this quantitative assessment. We believe that the threshold ratio of 1.5 is a useful parameter for the ATTR-CM diagnosis’ performance, both quantitatively and qualitatively.

## Data Availability

The datasets generated during and/or analyzed during the current study are not publicly available but are available from the corresponding author on reasonable request.

## Acknowledgments

Not applicable.

## Disclosures

### Authors’ contributions

All authors contributed to the study conception and design. Material preparation, data collection and analysis were performed by Julien Dubois, Florentin Kucharczak and Tom Paunet. The first draft of the manuscript was written by Julien Dubois and all authors edited and approved the manuscript.

